# Brain Activation in Chronic Nonspecific Low Back Pain : A Systematic review and ALE Meta-analysis

**DOI:** 10.1101/2021.08.26.21262683

**Authors:** Sandipan Hazra, Samantak Sahu, Prasun Priya Nayak, Koushik Sarkar, V Srikumar, Gita Handa

## Abstract

Pain, a protective mechanism turns into a pathologic response when it becomes chronic. Recent evidences are pointing towards neuroplastic brain changes as the primary factor for the persisting pain in chronic nonspecific low back pain (cLBP). To summarise the previous fMRI studies, a coordinate-based ALE meta-analysis of resting functional brain imaging studies is carried out to identify the clusters activated in the brain in cLBP.

Literature survey: ‘PubMed’, ‘Scopus’ and ‘Sleuth’ were searched for studies with resting functional whole-brain imaging in cLBP. Till October 2020; 258, 238, and 7 studies were found respectively after search. The activity pattern was documented in ‘without stimulation’ and ‘with stimulation’ groups. The risk of bias was assessed by Joanna Briggs Institute critical appraisal checklist for analytical cross-section studies. Total seven (224 cLBP patients, 110 activation foci) and six studies (106 cLBP patients, 66 activation foci) were selected among 277 studies for metanalysis in the ‘without stimulation’ and ‘with stimulation’ group respectively. In the ‘without stimulation’ group 8 statistically significant clusters were found. The clusters are distributed in the prefrontal cortex, primary somatosensory cortex, and primary motor cortex, anterior cingulate cortex, insular cortex, putamen, claustrum, amygdala, and associated white matters in both hemispheres. On the other group, 3 statistically significant clusters were found in the frontal cortex, Parietal cortex, and Insula. In the ‘with stimulation’ group, significant lateralization was observed and most of the clusters were in the right hemisphere. The white matter involvement was more in the ‘with stimulation’ group (78.62% Vs 38.21%). The statistically significant clusters found in this study indicate a probable imbalance in GABAergic modulation of brain circuit and dysfunction in descending pain modulation system. This disparity in pain neuro-matrix is the source of spontaneous and persisting pain in cLBP.

## Introduction

Pain, a cornerstone protective mechanism turns into a pathologic response when it becomes chronic. The relationship of tissue injury and nociception become dynamic and wrangled. The debate is how it becomes chronic and if it is chronic what will be the treatment strategy. From the time of French philosopher Renee Descartes (1644), pain is considered as physiologically specialized. The specific pain receptors in the body projects the information through nerve fibers in the specific regions in the brain (1). Alternatively, the recent demonstration of central sensitization is considered the pain can be maintained and modulated in brain (2). So, researchers tried to co-relate the pain in chronic non-specific low back pain (cLBP) with degenerative spinal changes and neuroplastic brain changes.

Kergel et al. (3) has pointed out neuroplastic changes in both grey and white matters in a Systematic review of functional brain imaging studies in cLBP patients. They also identified there is increased activation in few pain processing brain areas like, medial prefrontal cortex (medial PFC), cingulate cortex, amygdala and insula during rest. Increased activation was demonstrated in the medial PFC, cingulate cortex, amygdala, insula, primary somatosensory cortex (S1), primary motor cortex (M1) and secondary somatosensory cortex (S2) after painful stimuli or physical manoeuvres. In an another meta-analysis of 293 patients Yuan et al (4) has shown decrease in grey matter volume in bilateral medial prefrontal cortex (mPFC), anterior cingulate cortex and right orbitofrontal cortex. In another Systematic review of structural and functional brain changes in chronic low back pain, authors have suggested that brain changes corroborate brain emotional network rather than nociceptive pathway (5).

Approaches towards evaluation and treatment strategies of cLBP centred on peripheral spinal factors of cLBP has been challenged by these equivocal evidences. Recent evidences are pointing towards neuroplastic brain changes as primary factor for the persisting pain in cLBP. In the molecular and cellular level, chronic nociception leads to brain reorganization. Theses microscopic changes results in release of excitatory and inhibitory neurochemicals from neurones and glial cells, upregulation of ionotropic and metabotropic receptors by signalling pathways and alteration in presynaptic and postsynaptic neuronal excitability (6). The final outcome of these molecular changes are long-term potentiation and central sensitization (7)(8). These neuroplastic changes are reflected in altered functional brain connectivity. Therefore, there is an inevitable need to explore the mechanisms of pain in cLBP in order to optimize our diagnostic and therapeutic strategies.

In the clinical practice guideline chronic nonspecific low back pain is defined as *“pain occurring primarily in the back with no signs of a serious underlying condition (such as cancer, infection, or cauda equina syndrome), spinal stenosis or radiculopathy, or another specific spinal cause (such as vertebral compression fracture or ankylosing spondylitis)”* (9). Low back pain more than 3 months of duration is defined as chronic low back pain (9). Degenerative changes on imaging of lumber spine are usually considered nonspecific, as they correlate poorly with symptoms (9).

In this context the previous brain imaging studies in cLBP are summarised in this meta-analysis. The gold standard statistical analysis for imaging studies are meta-analysis from full statistical map of previously published studies by aggregating the effect size at each voxel (10). As full statistical maps are rarely available, peak co-ordinate-based methods are used commonly. This Co-ordinate based Activation likelihood estimation (ALE) analysis asses the consistency of activation in each voxel [unitary three dimensional (3D) point in 3D image, here in brain MRI]. ALE based meta-analysis of resting functional brain imaging studies is carried out to identify the areas activated in chronic nonspecific low back pain patients.

## Methodology

### Registration

The protocol of this review is registered in PROSPERO (registration number : CRD42020203007).

### Eligibility criteria

Studies with resting functional whole brain imaging in patients with chronic non-specific low back pain were included. Studies without brain imaging, reviews, animal studies, case reports, without full text, non-English language studies, without any peer review, not including whole brain analysis, with only structural imaging, studies not mentioning any standard stereotactic space 3 D coordinates x,y,z ; like Talairach or Montreal Neurological Institute for peak co-ordinate were excluded. The selected studies were done in resting conditions and with or without mechanical, thermal and pressure stimulations. Previous studies have shown that in chronic low back pain patients different brain areas are activated in resting fMRI with or without thermal stimulation (11) (12). Apkarian concluded that it is difficult to interpret the functional MRI studies with mechanical and thermal stimulation in spontaneous pain like CBP (13). So, we have categorised the fMRI studies in two groups, i.e., with stimulation and without stimulation. If any eligible article has reported fMRI data in both the condition, we have included the peak co-ordinate separately.

### Information source : Search and Data Item

Literature was searched online by using ‘PubMed’, ‘Scopus’ and ‘Sleuth’ (BrainMap database). We have also searched the reference list of relevant studies and review articles additionally. The main keywords used were (Chronic low back pain) AND (Brain OR Brain Activity OR Cortical changes OR Cortex OR Cortical activity OR Synapse OR Synaptic changes OR Sensorimotor processing OR Plasticity) AND (Central Nervous System Sensitization OR Sensitization OR Central sensitivity OR Central hyper-excitability OR Central sensitization OR Pain modulation OR Neural inhibition OR Hyperalgesia OR Nociception OR Pain threshold OR Algometry OR Hypersensitivity OR Gray matter OR White matter OR Functional connectivity) AND (MRI OR Magnetic resonance imaging OR fMRI OR Functional magnetic resonance imaging OR PET OR Positron emission tomography OR evoked potential OR NIRS OR fNIRS OR functional near-infrared spectroscopy OR Optical neuroimaging study OR Diffusion tensor imaging OR EEG OR Electroencephalography OR Brain imaging). Searching was done by SH & SS independently, and was supervised by GH & SV. All the authors has completed speciality training in Physical Medicine and Rehabilitation. They have 6, 4, 27 and 20 years of experiences in the speciality respectively.

### Study Selection

In ‘PubMed’, ‘Scopus’ and ‘Sleuth’ (BrainMap online database) we have found 258, 238 and 7 studies till October 2020 respetively. Additionally, after searching the reference list of relevant studies and review articles we have found additional ten references. After removing the duplicate studies, we have found 277 studies. Out of these, 120 studies were excluded for not having brain imaging studies, reviews, animal studies, case reports, non-availability of full text, non-English language studies, studies that are not peer reviewed. From that149 studies were excluded for not having whole brain analysis, reporting structural imaging only, not mentioning standard stereotactic space coordinates (Talairach or Montreal Neurological Institute), not satisfying the case definition of chronic low back pain (9). We have divided the studies full filling the inclusion criteria in two categories, i.e, (1) without lower back stimulation and (2) with lower back stimulation. In the first group one study by Baliki et al was included in qualitative analysis only (14). Because in this study the peak co-ordinates were not reported. Finally, seven studies were included in meta-analysis in the ‘without stimulation’ group. In the second group six studies were included for qualitative and quantitative analysis (figure 1).

**Figure : 1:**
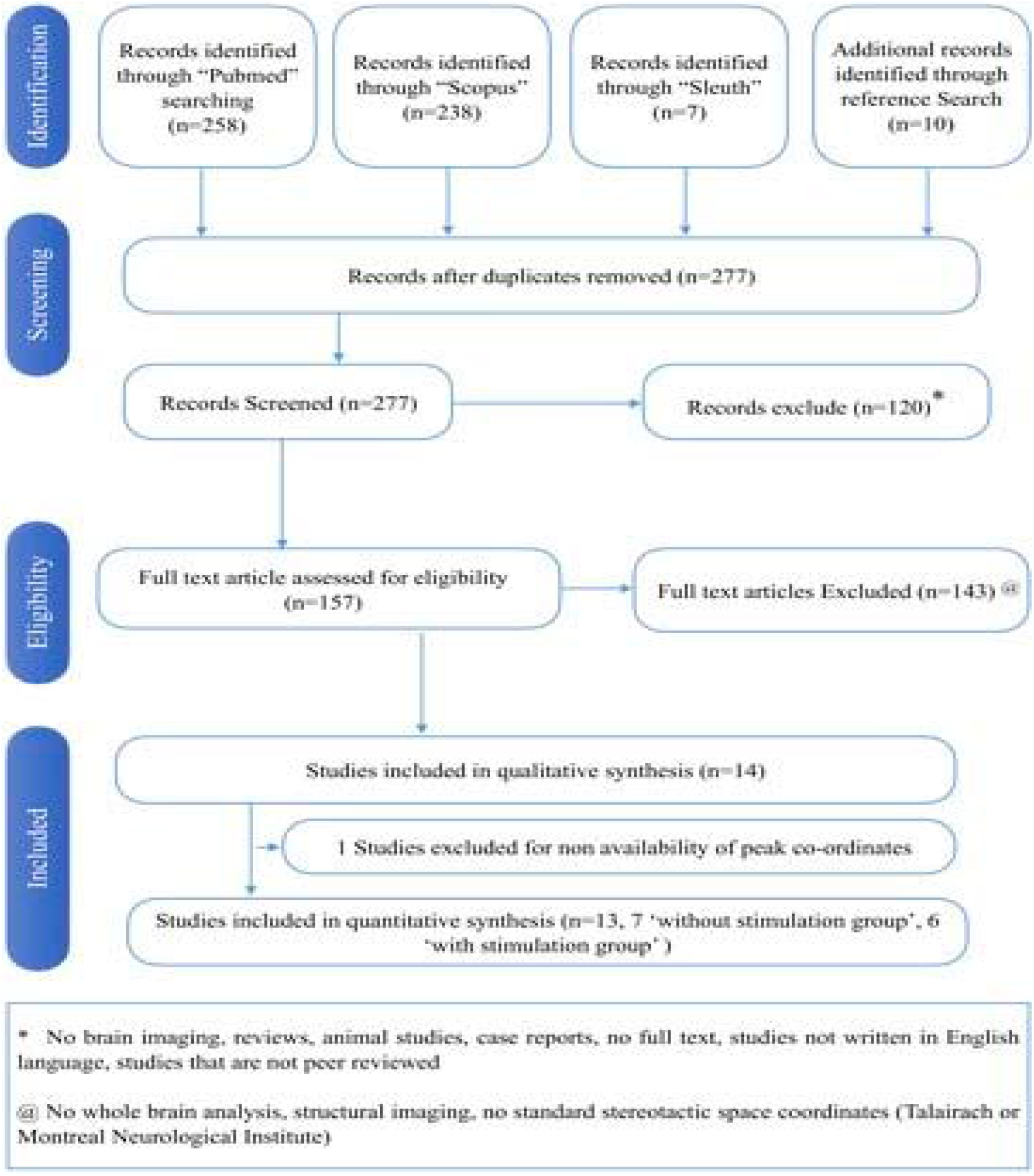
PRISMA FLOW Diagram showing the sequence of literature search and process of inclusion and exclusion of articles according to PRISMA statement (http://www.equator-network.org/reporting-guidelines/prisma/)

### Risk of bias assessment

Risk of bias was assessed by Joanna Briggs Institute critical appraisal check list for analytical cross-section studies, developed by Faculty of Health and Medical Sciences at the University of Adelaide, South Australia (15). It has eight items and each item have four options for answering. The answers are yes, no, unclear and not applicable.

### ALE Meta-analysis

ALE meta-analysis is summarising the co-ordinates in a voxel based analysis to know which regions are consistently activated (10). It is used to localise the pattern of anatomical brain region activated in a particular type of task. Null hypothesis for ALE method is that foci of activations are uniformly spread in the whole brain. So, this statistical method is used to assess the activation probabilities for each voxel in the brain. The null hypothesis is rejected when at least one peak co-ordinate falls within the voxels. In ALE, Monte Carlo procedure generates n number peaks at random locations. The peaks are assumed to follow a Gaussian distribution but the mean and variance are unknown. Primarily, a random mean and standard deviation is considered. Then the mean and deviation are adjusted to minimise the objective function. Doing so, a distribution is fitted, which is closely matched with the voxel distribution and they are grouped group on similarity measures (16). ALE value is calculated by union of peak probabilities where the probability is statistically significant. Z-score is used to standardise the distribution for comparison. Resultantly, Z-score have a distribution with a mean 0 and standard deviation 1. Accordingly, this signifies how far is the point from the mean of a data point. We have used GingerALE 3.0.2 for ALE meta-analysis and this is based on the protocol proposed by Eickhoff et al. (17) (18). For the threshold of ALE map, P value for the cluster-level family-wise error (FWE) was chosen <0.05 (corrected) and for cluster formation voxel-level forming threshold P value was chosen <0.001 (uncorrected) (19)(20). The p-value can be calculated as corrected and uncorrected. The observed p-value is also known as uncorrected p-value can be adjusted as suggested by Bonferroni is known as corrected p-value. We have chosen threshold for minimum cluster size as >200 mm^3^ (19). For visualization of results we have used Mango (4.1) by Research Imaging Institute, UTHSCSA and anatomical template provided on GingerALE Website (Colin27_T1_seg_MNI.nii, http://brainmap.org/ale). This template was overlaid with the ALE map generated by GingerALE 3.0.2. This ALE estimation and visualization was done by SH and KS.

## Results

### Study selection

Total 8 and 6 studies was selected among 277 studies for Systematic review for the ‘without stimulation’ and ‘with stimulation’ group respectively. Relevant information from the included studies were presented in the table 1a and 1b. The following items were included (1) study, (2) objective, (3) inclusion criteria, (4) exclusion criteria, (5) participants details and (6) location of brain where peak activity is reported. Most of the studies haave mentioned, the study population were right-handed.

**Table 1a:**
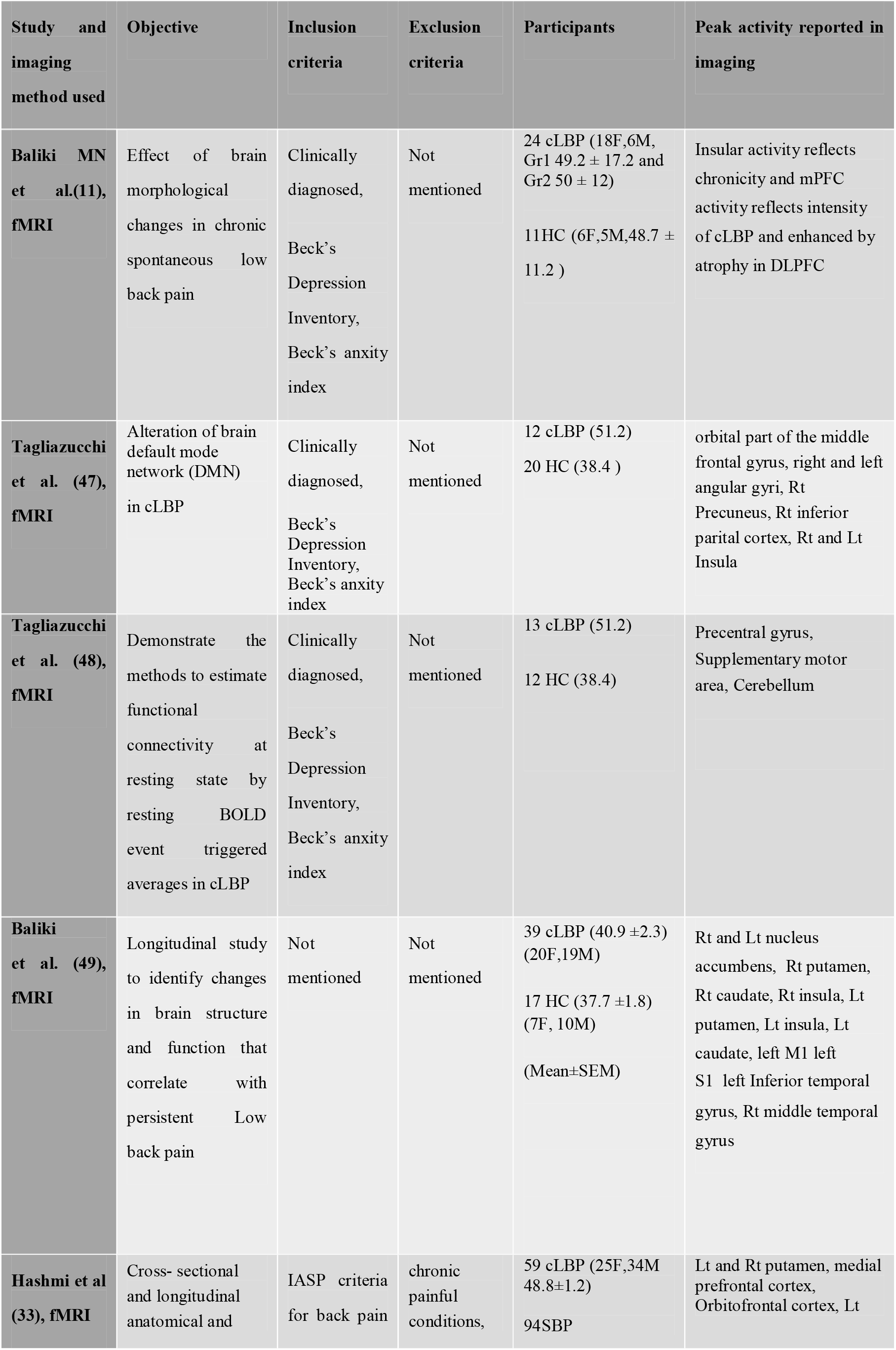
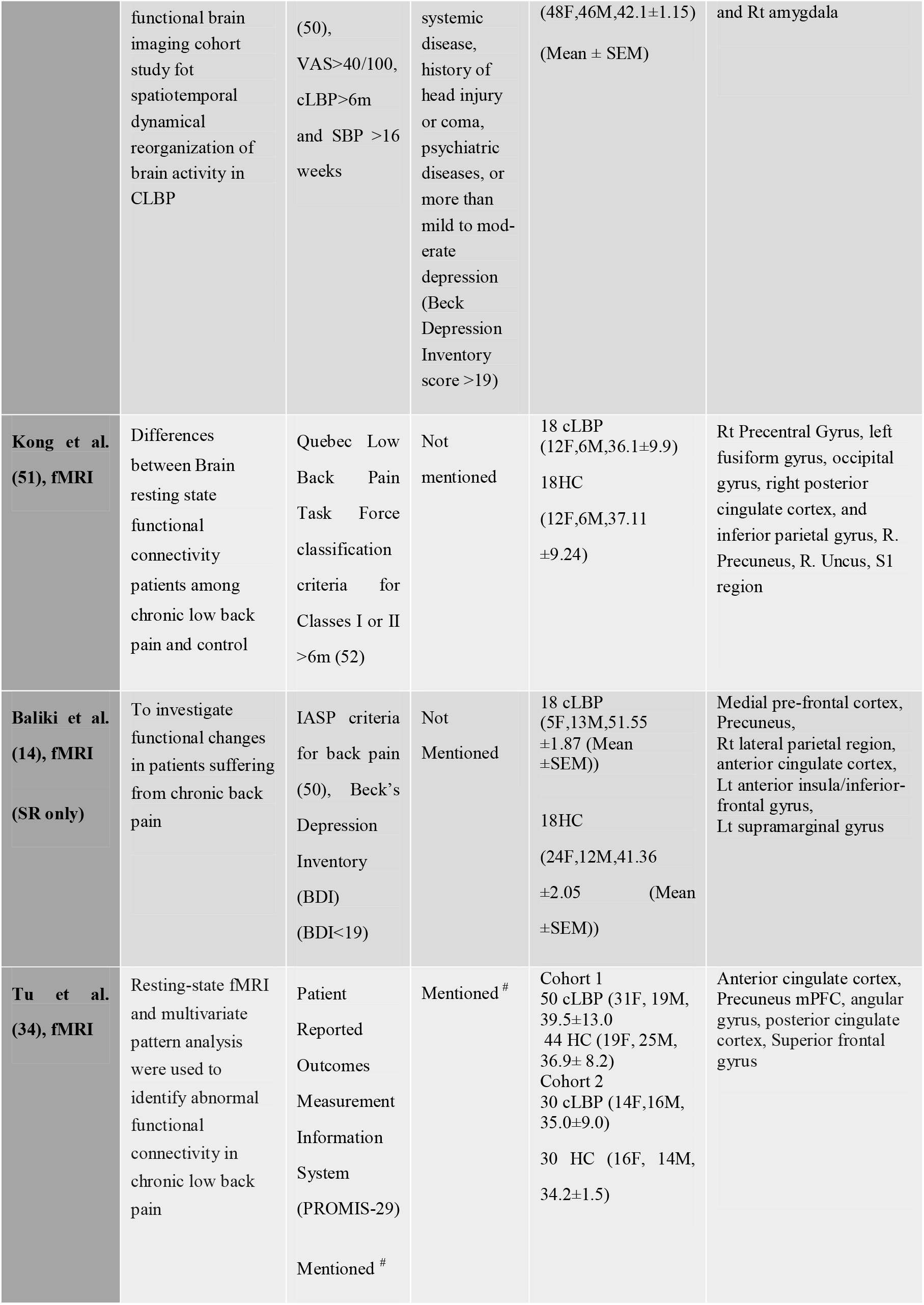
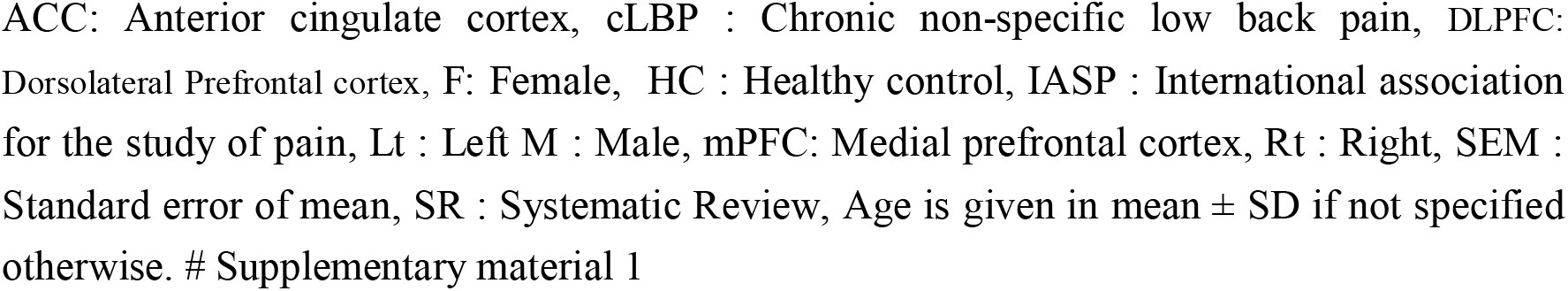
Summary of included fMRI studies in ‘without stimulation’ group.

Brain Activation in Chronic Nonspecific Low Back Pain : A Systematic review and ALE Meta-analysis
| Study and imaging method used | Objective | Inclusion criteria | Exclusion criteria | Participants | Peak activity reported in imaging |
| --- | --- | --- | --- | --- | --- |
| **Baliki MN et al.(11), fMRI** | Effect of brain morphological changes in chronic spontaneous low back pain | Clinically diagnosed, Beck's Depression Inventory, Beck's anxiety index | Not mentioned | 24 cLBP (18F,6M, Gr1 $49.2 \pm 17.2$ and Gr2 $50 \pm 12$)<br>11HC (6F,5M, $48.7 \pm 11.2$) | Insular activity reflects chronicity and mPFC activity reflects intensity of cLBP and enhanced by atrophy in DLPFC |
| **Tagliazucchi et al. (47), fMRI** | Alteration of brain default mode network (DMN) in cLBP | Clinically diagnosed, Beck's Depression Inventory, Beck's anxiety index | Not mentioned | 12 cLBP (51.2)<br>20 HC (38.4) | orbital part of the middle frontal gyrus, right and left angular gyri, Rt Precuneus, Rt inferior parital cortex, Rt and Lt Insula |
| **Tagliazucchi et al. (48), fMRI** | Demonstrate the methods to estimate functional connectivity at resting state by resting BOLD event triggered averages in cLBP | Clinically diagnosed, Beck's Depression Inventory, Beck's anxiety index | Not mentioned | 13 cLBP (51.2)<br>12 HC (38.4) | Precentral gyrus, Supplementary motor area, Cerebellum |
| **Baliki et al. (49), fMRI** | Longitudinal study to identify changes in brain structure and function that correlate with persistent Low back pain | Not mentioned | Not mentioned | 39 cLBP ($40.9 \pm 2.3$) (20F,19M)<br>17 HC ($37.7 \pm 1.8$) (7F, 10M)<br>(Mean±SEM) | Rt and Lt nucleus accumbens, Rt putamen, Rt caudate, Rt insula, Lt putamen, Lt insula, Lt caudate, left M1 left S1 left Inferior temporal gyrus, Rt middle temporal gyrus |
| **Hashmi et al (33), fMRI** | Cross- sectional and longitudinal anatomical and | IASP criteria for back pain | chronic painful conditions, | 59 cLBP (25F,34M $48.8 \pm 1.2$)<br>94SBP | Lt and Rt putamen, medial prefrontal cortex, Orbitofrontal cortex, Lt |
|  | functional brain imaging cohort study for spatiotemporal dynamical reorganization of brain activity in CLBP | (50),<br>VAS>40/100,<br>cLBP>6m<br>and SBP >16 weeks | systemic disease, history of head injury or coma, psychiatric diseases, or more than mild to moderate depression (Beck Depression Inventory score >19) | (48F,46M,42.1±1.15)<br>(Mean ± SEM) | and Rt amygdala |
| **Kong et al. (51), fMRI** | Differences between Brain resting state functional connectivity patients among chronic low back pain and control | Quebec Low Back Pain Task Force classification criteria for Classes I or II >6m (52) | Not mentioned | 18 cLBP (12F,6M,36.1±9.9)<br>18HC (12F,6M,37.11 ±9.24) | Rt Precentral Gyrus, left fusiform gyrus, occipital gyrus, right posterior cingulate cortex, and inferior parietal gyrus, R. Precuneus, R. Uncus, S1 region |
| **Baliki et al. (14), fMRI (SR only)** | To investigate functional changes in patients suffering from chronic back pain | IASP criteria for back pain (50), Beck's Depression Inventory (BDI) (BDI<19) | Not Mentioned | 18 cLBP (5F,13M,51.55 ±1.87 (Mean ±SEM))<br>18HC (24F,12M,41.36 ±2.05 (Mean ±SEM)) | Medial pre-frontal cortex, Precuneus, Rt lateral parietal region, anterior cingulate cortex, Lt anterior insula/inferior-frontal gyrus, Lt supramarginal gyrus |
| **Tu et al. (34), fMRI** | Resting-state fMRI and multivariate pattern analysis were used to identify abnormal functional connectivity in chronic low back pain | Patient Reported Outcomes Measurement Information System (PROMIS-29)<br>Mentioned # | Mentioned # | Cohort 1<br>50 cLBP (31F, 19M, 39.5±13.0)<br>44 HC (19F, 25M, 36.9± 8.2)<br>Cohort 2<br>30 cLBP (14F,16M, 35.0±9.0)<br>30 HC (16F, 14M, 34.2±1.5) | Anterior cingulate cortex, Precuneus mPFC, angular gyrus, posterior cingulate cortex, Superior frontal gyrus |

ACC: Anterior cingulate cortex, cLBP : Chronic non-specific low back pain, DLPFC: Dorsolateral Prefrontal cortex, F: Female, HC : Healthy control, IASP : International association for the study of pain, Lt : Left M : Male, mPFC: Medial prefrontal cortex, Rt : Right, SEM : Standard error of mean, SR : Systematic Review, Age is given in mean ± SD if not specified otherwise. # Supplementary material 1

**Table 1b:**
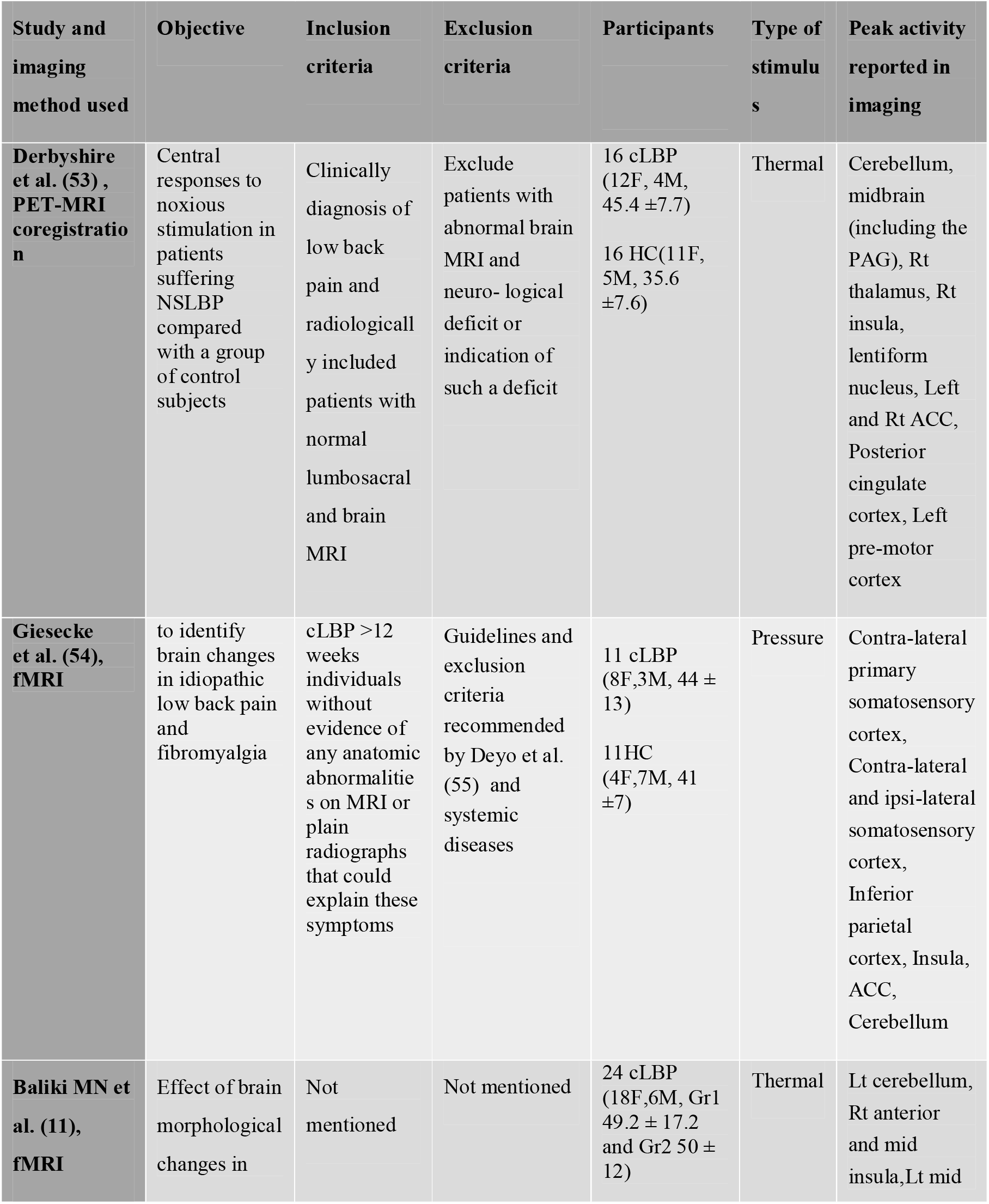
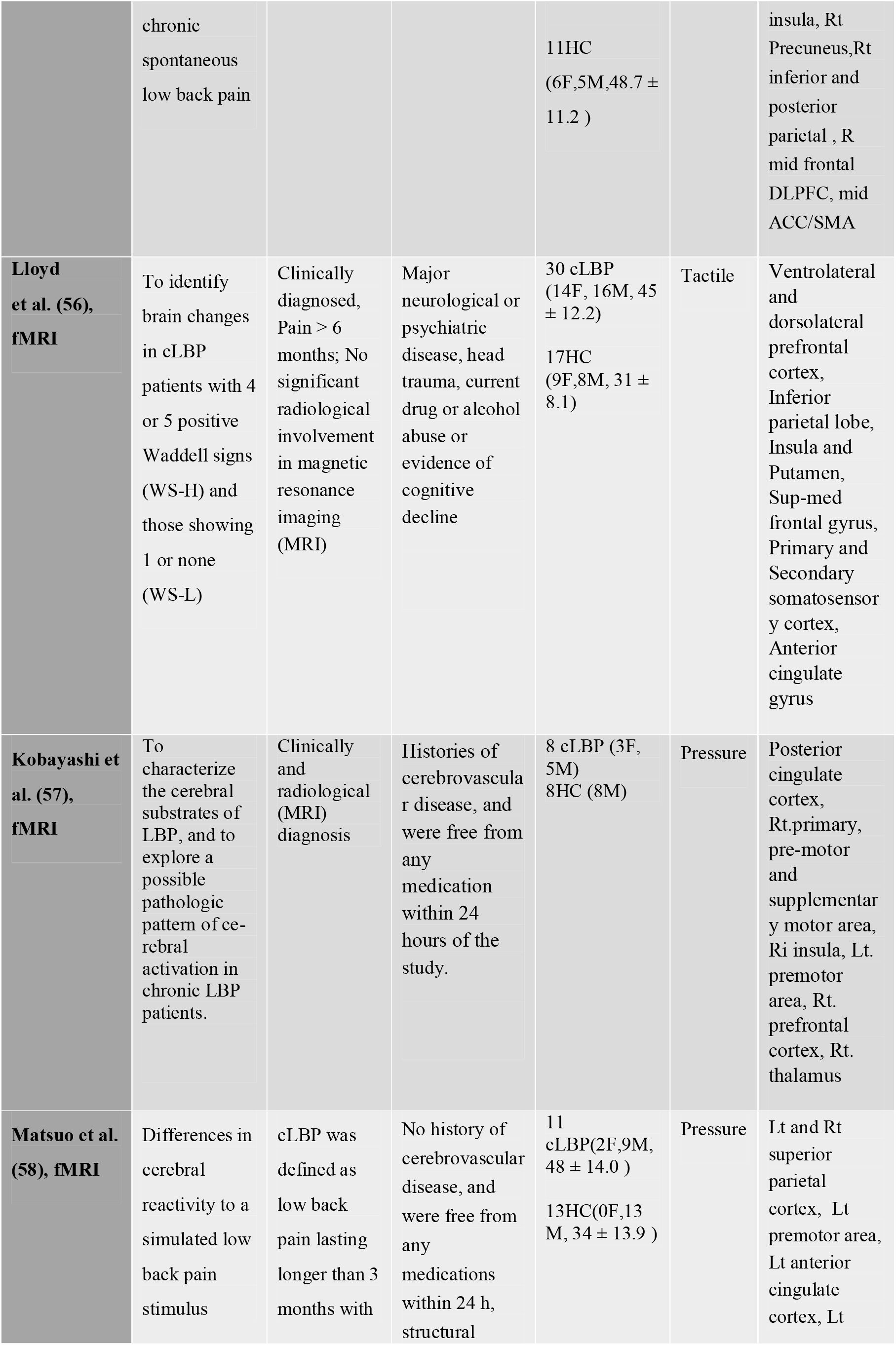
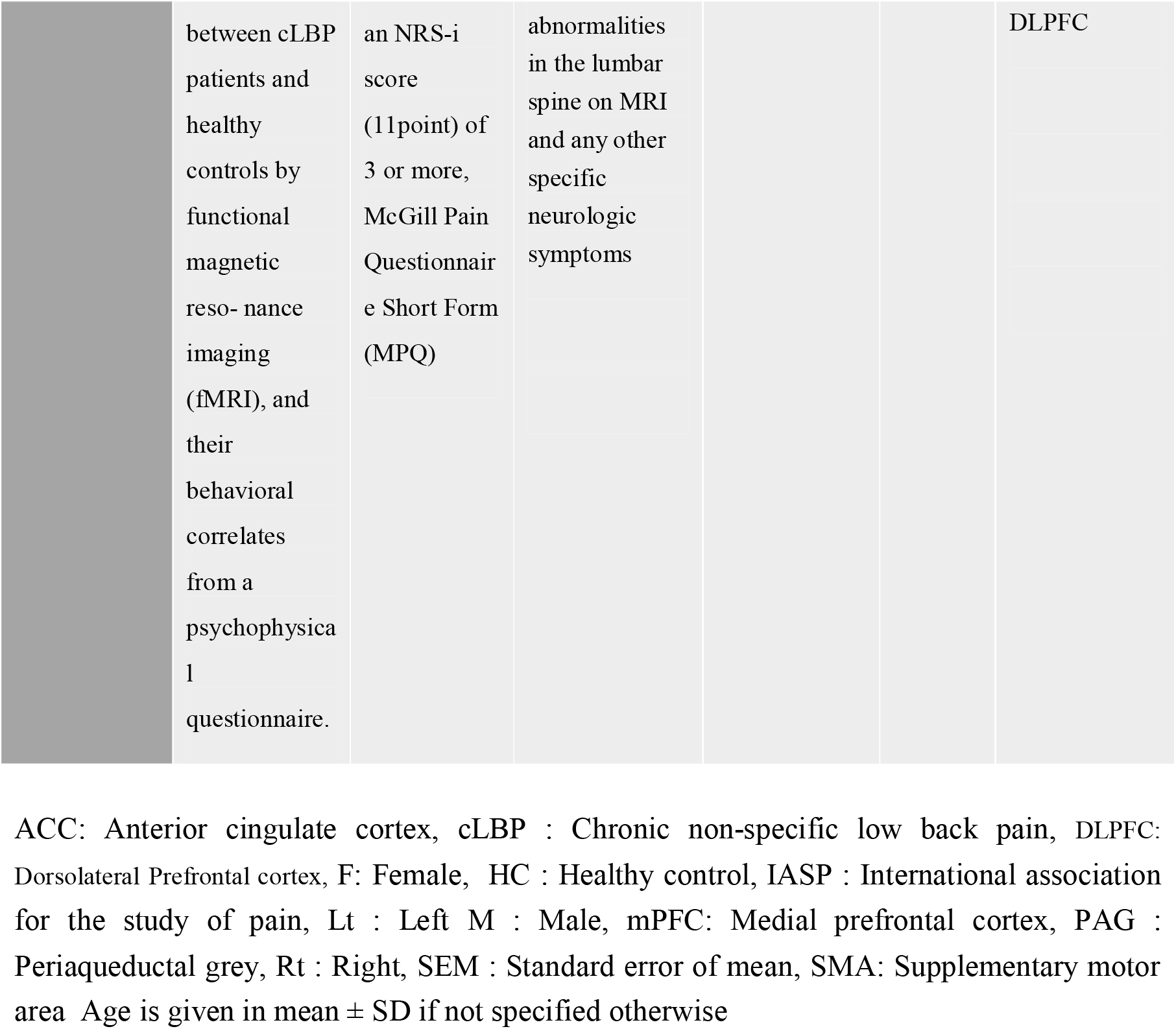
Summary of included fMRI studies in ‘with stimulation’ group.

### Risk of Bias

We have assessed the risk of bias of the 8 selected resting fMRI studies (table no 2a) and six fMRI studies with stimulation (table no 2b) in chronic non-specific low back pain by Joanna Briggs Institute critical appraisal check list.

**Table 2a:** Result of risk of bias assessment by Joanna Briggs Institute critical appraisal check list of the included studies in ‘without stimulation’ group.

| Author, date | Baliki et al.(11) | Tagliazucchi et al. (47) | Tagliazucchi et al. (48) | Baliki et al.(49) | Hashmi et al (33) | Kong et al. (51) | Baliki et al. (14) | Tu et al.(34) |
| --- | --- | --- | --- | --- | --- | --- | --- | --- |
| Were the criteria for inclusion in the sample clearly defined? | Unclear | Unclear | Unclear | Yes | Yes | Yes | Yes | Yes |
| Were the study subjects and the setting described in detail? | Yes | Yes | Yes | Unclear | Yes | Yes | Yes | Yes |
| Was the exposure measured in a valid and reliable way? | Yes | Yes | Yes | Yes | Yes | Yes | Yes | Yes |
| Were objective, standard criteria used for measurement of the condition? | No | No | No | No | Yes | Yes | Yes | Yes |
| Were confounding factors identified? | Unclear | No | No | Yes | Yes | No | Yes | Unclear |
| Were strategies to deal with confounding factors stated? | Unclear | No | No | Yes | Yes | No | Yes | No |
| Were the outcomes | Yes | Yes | Yes | Yes | Yes | Yes | Yes | Yes |
| measured in a valid and reliable way? |  |  |  |  |  |  |  |  |
| Was appropriate statistical analysis used? | Yes | Yes | Yes | Yes | Yes | Yes | Yes | Yes |

**Table 2b:** Result of risk of bias assessment by Joanna Briggs Institute critical appraisal check list of the included studies in ‘with stimulation’ group.

| Author, date | Derbyshire et al. (53) | Giesecke et al. (54) | Baliki et al. (11) | Lloyd et al.(56) | Kobayashi et al. (57) | Matsuo et al. (58) |
| --- | --- | --- | --- | --- | --- | --- |
| Were the criteria for inclusion in the sample clearly defined? | Yes | Yes | Unclear | Yes | Yes | Yes |
| Were the study subjects and the setting described in detail? | Yes | Yes | Yes | Yes | Yes | Yes |
| Was the exposure measured in a valid and reliable way? | Yes | Yes | Yes | Yes | Yes | Yes |
| Were objective, standard criteria used for measurement of the condition? | Unclear | Yes | No | Yes | Yes | Yes |
| Were confounding factors identified? | Yes | Yes | Unclear | Unclear | No | Unclear |
| Were strategies to deal with confounding factors stated? | Unclear | Yes | Unclear | Unclear | No | No |
| Were the outcomes measured in a valid and reliable way? | Yes | Yes | Yes | Yes | Yes | Yes |
| Was appropriate statistical analysis used? | Yes | Yes | Yes | Yes | Yes | Yes |

### ALE meta-analysis results

An ALE meta-analysis of total 7 and 6 selected studies was done in ‘without stimulation’ and ‘with stimulation’ group respectively. In the ‘without stimulation’ group, 110 activation foci was considered from a pooled data of 224 patients. In the ‘with stimulation’ group 66 activation was identified from a pooled data of 106 patients. Minimum cluster size was chosen as 200 mm^3^. Statistically significant clusters are described in the table 3a & 3b, and figure 2a,2b. In the ‘without stimulation’ group statistically significant activation found in the both hemispheres and frontal, parietal, limbic and sub-lobar regions. In the ‘with stimulation’ group a significant laterization was observed and most of the clusters were in right hemisphere (figure 2a,2b). Activation was observed in frontal lobe, parietal lobe, insula and limbic lobe. Both the grey and white matter areas are activated in both of groups. White matter involvement is more in fMRI studies with stimulation than without stimulation group (78.62% Vs 38.21%).

**Table 3a.** Results of ALE meta-analysis of resting fMRI studies ‘without stimulation’ group.

| Cluster | X | Y | Z | ALE (10<sup>-3</sup>) | Z Score | Size (mm<sup>3</sup>) | Label |
| --- | --- | --- | --- | --- | --- | --- | --- |
| 1 | -4 | 40 | -12 | 16.66 | 4.62 | 624 | **Hemisphere:** 76.9% Left Cerebrum, 16.7% Inter-Hemispheric, 6.4% Right Cerebrum<br>**Lobe:** 76.9% Limbic, 6.4% Frontal<br>**Gyrus:** 76.9% Anterior Cingulate, 6.4% Medial Frontal Gyrus<br>**Tissue Type:** 71.8% Gray Matter, 28.2% White Matter<br>**Cell Type:** 28.2% BA 32, 9% BA 24, 5.1% BA 10<br>**Contributing studies :** Baliki et al. |
|  |  |  |  |  |  |  | (11), Hashmi et al. (33), Tu et al. (34) |
| 2 | -42 | -20 | 56 | 15.00 | 4.30 | 464 | **Hemisphere:** 100% Left Cerebrum<br>**Lobe:** 72.4% Parietal, 27.6% Frontal<br>**Gyrus:** 62.1% Postcentral Gyrus, 37.9% Precentral Gyrus<br>**Tissue Type:** 84.5% Gray Matter, 15.5% White Matter<br>**Cell Type :** 37.9% BA 3, 25.9% BA 4<br>**Contributing studies :** Tagliazucchi et al. (47), Tagliazucchi et al. (48), Tu et al. (34) |
| 3 | -4 | 48 | 0 | 17.85 | 4.85 | 456 | **Hemisphere:** 96.5% Left Cerebrum, 3.5% Inter-Hemispheric<br>**Lobe:** 82.5% Limbic, 14% Frontal<br>**Gyrus:** 82.5% Anterior Cingulate, 14% Medial Frontal Gyrus<br>**Tissue Type:** 82.5% Gray Matter, 17.5% White Matter<br>**Cell Type :** 56.1% BA 32, 10.5% BA 10<br>**Contributing studies :** Hashmi et al. (33), Tu et al. (34) |
| 4 | -40 | -6 | -2 | 13.81 | 4.07 | 416 | **Hemisphere:** 100% Left Cerebrum<br>**Lobe:** 100% Sub-lobar<br>**Gyrus:** 75% Insula, 13.5% Extra-Nuclear, 11.5% Claustrum<br>**Tissue Type:** 69.2% White Matter, 30.8% Gray Matter<br>**Cell Type :** 19.2% Brodmann area 13<br>**Contributing studies :** Tagliazucchi et al. (48), Baliki et al. (49), Kong et al. (51) |
| 5 | -14 | 56 | 20 | 14.06 | 4.12 | 328 | **Hemisphere:** 100% Left Cerebrum<br>**Lobe:** 100% Frontal Lobe<br>**Gyrus:** 100% Superior Frontal Gyrus<br>**Tissue Type:** 90.2% White Matter, 9.8% Gray Matter<br>**Cell Type :** 9.8% Brodmann area 9<br>**Contributing studies :** Baliki et al. (11), Tu et al. (34). |
| 6 | -28 | 8 | 0 | 11.93 | 3.74 | 264 | **Hemisphere:** 100% Left Cerebrum<br>**Lobe:** 100% Sub-lobar<br>**Gyrus:** 57.6% Lentiform Nucleus, 42.4% Extra-Nuclear<br>**Tissue Type:** 57.6% Gray Matter, 42.4% White Matter<br>**Cell Type:** 57.6% Putamen<br>**Contributing studies:** Hashmi et al. (33), Tu et al. (34) |
| 7 | -24 | -4 | -18 | 11.94 | 3.74 | 224 | **Hemisphere:** 100% Left Cerebrum<br>**Lobe:** 100% Limbic Lobe<br>**Gyrus:** 100% Parahippocampal Gyrus<br>**Tissue Type:** 67.9% Gray Matter, 32.1% White Matter<br>**Cell Type:** 67.9% Amygdala<br>**Contributing studies:** Kong et al. (51), Hashmi et al. (33) |
| 8 | 50 | -12 | 52 | 12.68 | 3.88 | 216 | **Hemisphere:** 100% Right Cerebrum<br>**Lobe:** 70.4% Parietal Lobe, 29.6% Frontal Lobe<br>**Gyrus:** 66.7% Postcentral Gyrus, 33.3% Precentral Gyrus<br>**Tissue Type:** 77.8% Gray Matter, 22.2% White Matter<br>**Cell Type:** 55.6% BA 3, 22.2% BA 4<br>**Contributing studies:** Tagliazucchi et al. (47), Tagliazucchi et al. (48) |

**Table 3b.** Results of ALE meta-analysis of resting fMRI studies ‘with stimulation’ group.

| Cluster | X | Y | Z | ALE<br>($10^{-3}$) | Z<br>Score | Size<br>($\text{mm}^3$) | Label |
| --- | --- | --- | --- | --- | --- | --- | --- |
| 1 | 50 | 4 | 10 | 18.92 | 5.61 | 1560 | **Hemisphere:** 100% Right Cerebrum<br>**Lobe:** 53.3% Frontal Lobe, 46.7% Sub-lobar<br>**Gyrus:** 45.6% Insula, 40% Precentral Gyrus, 13.3% Sub-Gyral, 1% Extra-Nuclear<br>**Tissue Type:** 78.5% White Matter, 21.5% Gray Matter<br>**Cell Type:** 11.8% BA 44, 9.7% BA 13<br>**Contributing studies:** Derbyshire et al. (53), Baliki et al. (11), Lloyd et al. (56), Kobayashi et al. (57) |
| 2 | 62 | -20 | 18 | 15.34 | 4.87 | 576 | **Hemisphere:** 100% Right Cerebrum<br>**Lobe:** 100% Parietal Lobe<br>**Gyrus:** 100% Postcentral Gyrus<br>**Tissue Type:** 88.9% White Matter, 11.1% Gray Matter<br>**Cell Type:** 9.7% BA 40<br>**Contributing studies:** Giesecke et al. (54), Loyd et al. (56) |
| 3 | 4 | 20 | 42 | 11.70 | 4.15 | 480 | **Hemisphere:** 61.7% Right Cerebrum, 23.3% Inter-Hemispheric, 15% Left Cerebrum<br>**Lobe:** 50% Frontal Lobe, 26.7% Limbic Lobe<br>**Gyrus:** 26.7% Cingulate Gyrus, 26.7% Medial Frontal Gyrus, 23.3% Superior Frontal Gyrus<br>**Tissue Type:** 66.7 White Matter, 33.3% Gray Matter<br>**Cell Type:** 15% BA 8, 10% BA 6, 8.3% BA 32<br>**Contributing studies:** Giesecke et al. (54), Lloyd et al. 2008 (56), Kobayashi et al. (57) |

**Figure : 2a:**
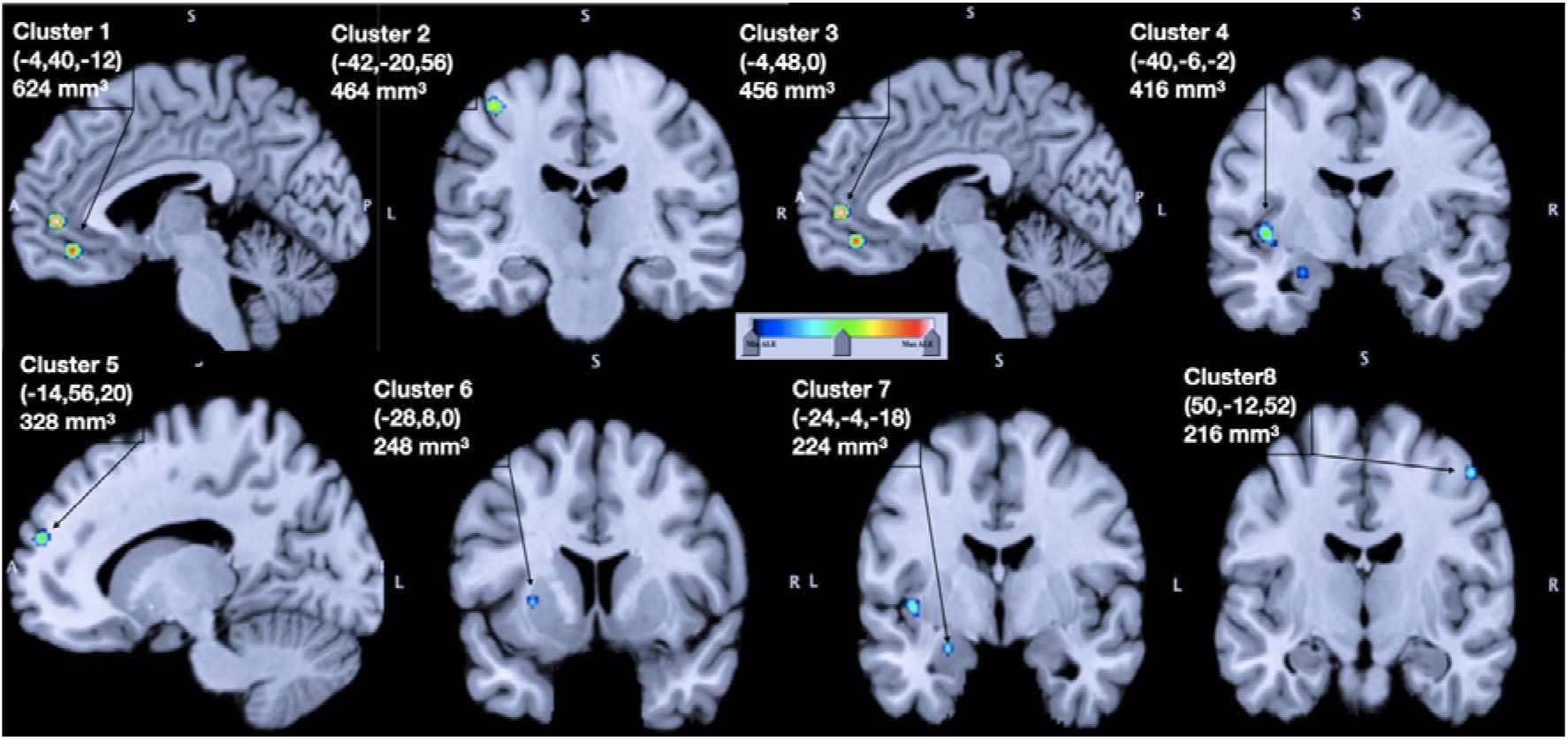
Result of ALE meta-analysis for the studies in ‘without stimulation’ group. The result is overlaid on the brain anatomical template provided on GingerALE Website (Colin27_T1_seg_MNI.nii, http://brainmap.org/ale). Red : Strong association, Green : Moderate association, Blue : Weaker association. Co-ordinates of the clusters (X,Y,Z) are provided in MNI (Montreal Neurological Institute) co-ordinate System of the human brain, mm^3^ : area of the cluster in cubic millimetre. Cluster forming values : threshold of ALE map the P <0.05 corrected for cluster-level family-wise error (FWE) and P<0.001 voxel-level forming threshold (uncorrected), threshold for minimum cluster size is >200 mm^3^

**Figure : 2b :**
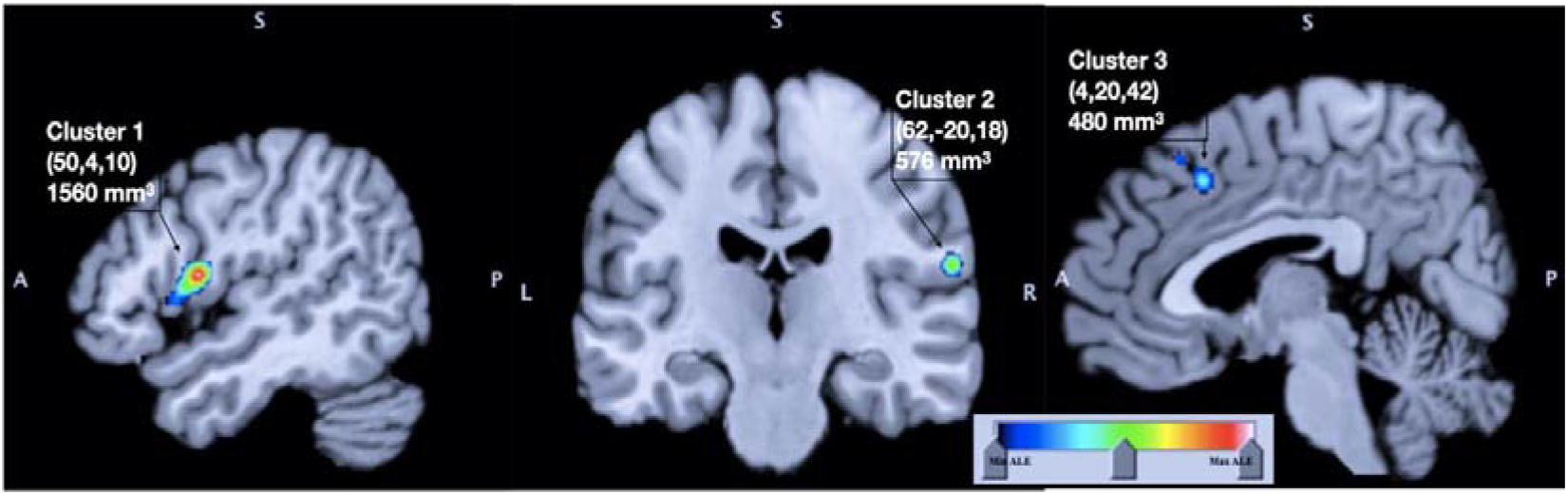
Result of ALE meta-analysis for the studies in ‘with stimulation’ group. The result is overlaid on the brain anatomical template provided on GingerALE Website (Colin27_T1_seg_MNI.nii, http://brainmap.org/ale). Red : Strong association, Green : Moderate association, Blue : Weaker association. Co-ordinates of the clusters (X,Y,Z) are provided in MNI (Montreal Neurological Institute) co-ordinate System of the human brain, mm^3^ : area of the cluster in cubic millimetre. Cluster forming values : threshold of ALE map the P <0.05 corrected for cluster-level family-wise error (FWE) and P<0.001 voxel-level forming threshold (uncorrected), threshold for minimum cluster size is >200 mm^3^

## Discussion

The fMRI assess spatial activity in the entire brain at rest in ‘with stimulation’ or ‘without stimulation’ group in cLBP patients. In fMRI BOLD MRI signals reflect the neuronal activity based on the neuronal oxygenation. The activity pattern was different in spontaneous pain and hyperalgesia – allodynia. This is reflected in the finding of ‘without stimulation’ and ‘with stimulation’ group respectively. In this ALE meta-analysis of seven fMRI ‘without stimulation’ studies, we have found eight statistically significant clusters activated in ‘spontaneous’ chronic non-specific low back pain. Those are distributed in prefrontal-cortex, primary somatosensory cortex and primary motor cortex, anterior cingulate cortex, insular cortex, putamen, claustrum, amygdala and associated white matters (table 3a, and figure 2a). Five studies have mentioned the diagnostic criteria they have used to define cases (table 2a). Other studies included the patients diagnosed clinically among them one study did not mention the criteria. Five studies have mentioned about the confounding factors.

We have found three statistically significant clusters in ALE meta-analysis of six fMRI studies of ‘with stimulation’ group in chronic non-specific low back pain or in ‘hyperalgesia – allodynia’. Those are in frontal cortex, parietal cortex and insula (table 3b, and figure 2b). In this group, five studies have mentioned the diagnostic criteria for case definition and only one study has specified about the confounding factors (table 2b). Significance of this identified clusters in cLBP is discussed in the following sections.

Pain is a multidimensional experience. According to pain matrix in the brain, four distinct regions can be identified to represents the facets of pain sensation - (a) sensory motor region, (b) cognitive regions, (c) affective regions, and (d) modulatory regions [5].

a. **Sensory - Motor region** The sensory discriminative element of pain indicates the intensity i.e., when, where and how (22). Along with that, primary somatosensory cortex (S1) processes some epicritic information and secondary somatosensory cortex (S2) deals with some higher cognitive component of pain (23). Primary motor cortex involvement in the chronic pain is associated with execution of movement. In a recent meta-analysis, Chang et al. has reported inconsistent association between those (24). There are evidences of expansion and shift of cortical areas in S1, S2, M1, ACC and insular cortex with chronicity of pain (25). In the ‘without stimulation’ group of this study, two statistically significant clusters are found; one in right and left primary somatosensory (BA3) and the other one in primary motor cortex (BA4) (table 3a and figure 2a). Similarly, another cluster is found right S2 cortex (BA40) of the ‘with stimulation’ group (table 3b). In this context, it is noteworthy that the insular and S1 cortex define the laterality of pain (26). The involvement of S2 cortex in the present study ‘with stimulation’ group possibly indicates the involvement of higher cognitive component in the case of hyperalgesia or allodynia and affective assignment of pain. Godinho et al. have concluded that a relatively late occurring responses in right somatosensory, temporo-occipital and temporal hemisphere are associated with memory encoding and emotional component of pain (27). From the present analysis, one cluster was also found in supplementary motor cortex (Rt BA6) and motor association cortex (Rt BA8) in the ‘with stimulation’ group (table 3b, and figure 2b). Mishra et al. have shown that these areas are connected with pain and motor control (28).
b. **Cognitive region:** Prefrontal activity is correlated with cognitive domain of nociception; therefore it is related with memory, attention, knowledge and understanding (23). However, Coghill et al. (29) and Strigo et al. (30) have shown that this area is not directly related with sensation and affect. Prefrontal cortex (BA 10,9) is associated with modulating nociception in “top-down” approach. More precisely, orbital frontal cortex (BA10) controls the affective perception (31). Several human and animal studies have documented time dependent decrease of grey matter volume in prefrontal cortex (32). Among the selected studies for the current analysis, there are three statistically significant clusters in pre-frontal cortex, ie, left anterior prefrontal cortex (BA 10), left dorsolateral and medial prefrontal cortex (BA 9) (table 3a and figure 2a) in the ‘without stimulation’ group. On the contrary, we have found no significant clusters of activation in the ‘with stimulation’ group (table 3b). Baliki et al. have also documented increased activity in medial prefrontal cortex (mPFC) (BA12, 24, 25,32, 33) and this activity was increased by atrophy of dorsolateral prefrontal cortex (BA8,9,10,46) (11). Hashmi et al. also pointed out increased activity in the mPFC amygdala and basal ganglia as a marker of cLBP (33). Further supporting these observations, the mPFC and rostral anterior cingulate cortex (rACC) have been found to have increased activity in a recent study by Tu et al.(34). In a Systematic review Kregel et al. also supports the involvement of pre frontal cortex in chronic low back pain (3).
c. **Affective region :** Affective component denotes the ‘unpleasantness’ of pain perception. Activation of cortico-limbic circuitry has been postulated as a risk-factor for development of chronic pain (23). In this study, the activation is found in left and right rostral anterior cingulate cortex (rACC) (BA32, BA24), left amygdala, left anterior insula and left basal ganglia. It is evident that activation of cortico-limbic pathway is more than that of mesolimbic pathway as there are no statistically significant activation of the regions like hippocampas, thalamus and mid brain. Among the selected studies in the ‘without stimulation’ group, activation of left rACC [dorsal anterior cingulate cortex (ACC) (BA 32) and ventral anterior cingulate cortex (BA 24)] (table 3a and figure 2a) was documented by Baliki et al. (11), Hashmi et al (33) and Tu et al.(34). Tolle et al (35) and Zubieta et al (36) also have shown ACC is more related with affective component of pain. Insular cortex (IC) has been found to be involved in sensory and affective dimensions of pain. Anterior insula performs as an integration site for multimodal information of pain including attention, anticipation and belief (23). It is also associated with pain intensity and and possible pain amplification (37). It has circuitry connection with PFC, ACC, amygdala and descending pain modulation system. Initially the glutaminergic receptors plays role in chronification of pain in IC. Subsequently, the imbalance between the glutaminergic, GABAergic and dopaminergic pathways contributing to the dysfunction in the pain processing and modulation (38). In both ‘with stimulation’ and ‘without stimulation’ groups, one cluster is found in insular cortex (BA13) in right and left lobe respectively (table 3a & 3b and figure 2a & 2b). Contralateral involvement of insula in the with stimulation group may be due to laterality of nociception as described in the previous section. In the pain processing, circuitries of central amygdala (CaA) and basolateral amygdala (BLA) complex act in conjunction. The CaA is associated with negative emotional aspects of pain and named as ‘nociceptive amygdala’(39). Whereas, BLA integrates polymodal sensory information (both noxious and non-noxious) and generate a memory regarding nociception (23). Amygdala acts as a central component of GABAergic circuitry in brain and controls the reward-aversion circuitry. It receives sensory information from spinal lamina 1 through spino-reticular pathway (parabrachial neucleus). Amygdala sends information to PFC and ACC and control the descending pain modulating system by periaqueductal grey (PAG) and rostral ventrolateral medulla (RVM) (23). In this meta-analysis, one statistically significant cluster is found in this area in ‘without stimulation’ group (table 3a, figure 2a). In the present analysis, two clusters are noted in the basal ganglia (Cluster 6, putamen and Cluster 4 claustrum) (table 3a, figure 2a). Basal ganglia is associated with motor, associative and emotional processing of pain (40). Putamen, in particular, is concerned in maintaining somatotopic map of pain (41) and subjective rating of pain (42).
d. **Modulatory region :** Electrical stimulation to peri-aqueductal grey (PAG) produces anti-nociceptive effect in dorsal horn spinal cord circuitry. The PAG output pathway is under influences of PFC and ACC and this connection is predominantly *GABaergic*. In cLBP patients this descending circuitry to PAG displays abnormal functional connectivity (43). The PAG-Spinal cord projection routes through rostral ventrolateral medulla (RMV). The PFC also acts as a connecting node in BLA-PFC-PAG circuitry. Prefrontal deactivation depresses this antinociceptive descending pathway. The final outcome is increased descending facilitation and decreased inhibition (pro-nociceptive) of PAG-RMV-spinal cord circuit (23). This pro-nociceptive priming of RMV causes reduction of threshold of activation of both ON and OFF cells to innocuous stimuli in chronic pain. The failure of compensatory rebalance and decreased top down modulation make the circuitry nocifensive (44). In this present study no significant cluster is identified in the descending pain modulatory region in either of the study groups (table 3a & 3b). This is probably due to decrease in PFC-PAG output.

The white matter involvement is greater in the ‘with stimulation’ group than the other one (78.62% Vs 38.21%) (table 3a and 3b). The chronic pain is associated with both ‘neuropathy’ and ‘gliopathy’. After injury, glial modulators causes activation of microglia and astrocytes. The activated glial cells release various neuromodulators and ultimately these induces synaptic and neuronal plasticity (45) (46). Perhaps the increase in white matter involvement in ‘with stimulation’ group is because of central sensitivity.

In this study, a stringent case definition of cLBP was followed to exclude the heterogenicity and ambiguity of case definition. This excludes few experiments included in the previous Systematic reviews. This face-off between power and homogeneity reduces the power of this study and this may affect the generalisability of the result (20). The data from the fMRI studies was extracted manually and was double checked by SH and KS to minimise the chances of error. The selected studies were of cross sectional study design with mode. Few studies didn’t mention about the diagnostic criteria and the confounding factors. Future meta-analysis with combining all the spatio-temporal domain of fMRI studies could be interesting. Additionally, inclusion of more with specific case definition in cLBP can increase the meta-analysis.

The nodes for the circuits are S1 cortex, rostral ACC (BA 32,24), prelimbic PFC (BA 10,9) and amygdala. Basal ganglia (putamen and claustrum) and M1 cortex activation are also noted along with these areas. These areas are involved in the cognitive, affective and sensory discriminative-efferent responses to pain. Nevertheless, no statistically significant activation is found in infra-limbic area (BA25), periaqueductal, rostral ventral medullary area and parabrachial area. These areas are associated with descending pain modulation system. There is no significant cluster found in the descending pain modulatory region in this meta-analysis. Hence, the fine-tuning balance between descending facilitation and inhibition in this circuit is altered. The resultant dysfunction in recruitment of descending pain modulation system create the circuit pro-nociceptive.

This meta-analysis of resting fMRI studies identified the statistically significant activation clusters in spontaneous pain and hyperalgesia – allodynia in cLBP patients. No activation is found in PAG-RMV-Spinal cord axis. Possibly the imbalance in GABAergic circuitry leads to dysfunction of descending pain modulation system and this altered pain neuro-matrix is the key mechanism for the persisting pain in cLBP.

## Data Availability

The data of meta-analysis is available in the public domain in the respective articles . GingerALE 3.0.2 was used for ALE meta-analysis and Mango (4.1) by Research Imaging Institute, UTHSCSA was used for visualization of the data.

